# Seroprevalence of hepatitis A and hepatitis E viruses among pregnant women in Northern Iran

**DOI:** 10.1101/2021.04.15.21255531

**Authors:** Farzin Sadeghi, Zahra Golchob, Maryam Javadian, Mohammad Barary, Parisa Sabbagh, Soheil Ebrahimpour, Masoumeh Bayani

## Abstract

**Background:** Hepatitis A (HAV) and Hepatitis E viruses (HEV) are endemic in Iran and are known major causes of acute viral hepatitis. Also, during pregnancy, they are associated with severe outcomes. Therefore, it is vital to evaluate the antibody levels against HAV and HEV in pregnant women to avoid severe outcomes incidence.

**Study design and methods:** A total of 247 pregnant women were enrolled in this prospective cross-sectional study. In addition to completing the questionnaire and interviewing all participants, the serum samples were tested for anti-HAV and anti-HEV IgG using the enzyme-linked immunosorbent assay (ELISA). The association between anti-HAV and anti-HEV antibodies status and risk factors was evaluated.

**Results:** The mean age of patients was 28.06 ± 5.29 years. Anti-HAV antibody was found in 111 patients (44.9%), while anti-HEV antibody was detected in only two pregnant women (0.8%). The seroprevalence of HAV was inversely related to the level of education. There was no significant correlation between HAV antibody levels and age, marital status, residence location, and pregnancy trimesters.

**Conclusion:** Considering many complications of these diseases in pregnancy, the detection of enteroviral hepatitis, especially HAV in pregnant women, is necessary, and therefore, proactive measures, such as promoting education, improving people awareness, and vaccination, are recommended

## Introduction

The principal causes of viral hepatitis are hepatitis A, B, C, and E viruses [1-4], among which the hepatitis A virus (HAV) and the hepatitis E virus (HEV) cause self-limiting acute hepatitis. These viruses spread via the fecal-oral route or by ingestion of contaminated water or food [5]. In many developing countries, these infections’ prevalence is closely associated with poor hygiene conditions and low socioeconomic status. In recent years, the incidence of HAV and HEV has decreased markedly due to improved health conditions. The risk of infection in youth was more significant than in adults [6]. Also, there was a high risk of vertical transmission of these viruses from the mother to the fetus with subsequent maternal and fetal complications, such as abortion, neonatal death, and premature labor [7, 8]. Thus, the diagnosis of these infections in pregnant women is vital to avoid any adverse outcomes. Laboratory results indicating an increase in anti-HAV or anti-HEV IgG levels in serum indicate past exposure or acute infection by these infections.

The levels of IgG antibodies in some asymptomatic populations related to HAV and HEV infections vary significantly. These levels are particularly variable among asymptomatic pregnant women. Besides, there are numerous reports that Iran is located in an endemic area of HAV infection, and several studies in Iran have shown that the HAV seroconversion rates are high among young Iranians [9]. Also, some reports indicated that the overall seroprevalence of HEV in Iran ranged from 2% to more than 40% [10].

With all this said, this study aimed to measure the seroprevalence of anti-HAV and anti-HEV antibodies and potential risk factors among pregnant women referred to Ayatollah Rouhani Hospital in Babol, Northern Iran.

## Materials and Methods

### Patients

This prospective cross-sectional study was comprised of 247 pregnant women aged 17-42 years. Pregnant women with no history of immunosuppressive diseases, such as human immunodeficiency viruses (HIV), cancers, chronic liver diseases, and use of corticosteroids, who were referred to Ayatollah Rouhani Hospital in Babol, Northern Iran, for their standard check-up services were enrolled. A standardized questionnaire was used to collect information on social, demographic data (age, education, place of residence), parity, and pregnancy trimesters. Before participating, the study’s purpose was clearly explained for all pregnant women, and written informed consent was obtained from all individuals.

### Laboratory analysis

Blood samples (5 mL) were collected from each participant, centrifuged, and separated serum was stored at -20^°^C until further analysis. It is noteworthy that all serum samples were duplicated for anti-HAV and anti-HEV total (IgG) antibodies analysis using an enzyme-linked immunosorbent assay (ELISA) commercial Kit (Dia.Pro Diagnostic Bioprobes Srl, Milan, Italy). For the anti-HEV IgG ELISA kit, the limit of detection and sensitivity was < 0.1 IU/mL and 0.25 IU/mL, respectively. Regarding the anti-HAV IgG ELISA kit, these characteristics were < 0.01 IU/mL and 0.15 IU/mL.

### Statistical analysis

Statistical analysis was performed using the SPSS software v. 16.0 (IBM, Chicago, IL, USA). A Chi-square test was used to compare anti-HEV/HAV seroprevalence for the mentioned stratification factors. A p-value < 0.05 was considered to be statistically significant.

## Results

Table 1 presents the demographic data of the study participants. Of the 247 pregnant women, 111 (44.9%) were positive for total anti-HAV IgG, and 2 (0.8%) were positive for anti-HEV IgG antibody. The overall mean age of pregnant women was 28.06 ± 5.29 years, with a range of 17-42 years. Besides, the mean ages of positive anti-HAV and anti-HEV antibodies were 27.9 ± 5.18 and 28.25 ± 5.43 years, respectively. Also, there was a strong inverse statistical association between education status and seroprevalence of HAV (p = 0.003). Our data did not show a statistically significant association between HAV infection and age, parity, place of residence, or pregnancy quarters (Table 2).

**Table 1.** Demographic characteristics of study patients.

| Demographic characteristics | Frequency<br>n (%) |
| --- | --- |
| **Age** |  |
| ≤ 30 | 148 (60) |
| > 30 | 99 (40) |
| **Educational status** |  |
| No formal education | 169 (68.4) |
| College and above | 78 (31.6) |
| **Place of residence** |  |
| Rural | 149 (60.3) |
| Urban | 98 (39.7) |
| **Parity** |  |
| First pregnancy | 123 (49.8) |
| Second | 107 (43.3) |
| Third or more | 17 (6.9) |
| **Pregnancy trimesters** |  |
| First | 69 (27.9) |
| Second | 98 (39.7) |
| Third | 80 (32.4) |

**Table 2.** Comparison of demographic characteristics of pregnant women between positive and negative anti-hepatitis A virus (HAV)

| Characteristics |  | HAV |  | Adjusted OR<br>(CI 95%) | p-value | Unadjusted OR<br>(CI 95%) | p-value |
| --- | --- | --- | --- | --- | --- | --- | --- |
|  |  | Negative | Positive |  |  |  |  |
| **Educational status** | No formal education | 84 (61/8) | 85 (76/6) | 1 | 0.003 | 1 | < 0.01 |
|  | College and above | 52 (38/2) | 26 (23/4) | 0.39<br>(0.21-0.72) |  | 0.49<br>(0.28-0.86) |  |
| **Place of residence** | Rural | 84 (61/8) | 65 (58/6) | 1 | 0.22 | 1 | 0.60 |
|  | Urban | 52 (38/2) | 46 (41/4) | 0.68<br>(0.36-1.26) |  | 0.87<br>(0.52-1.46) |  |
| **Parity** | First pregnancy | 70 (51/5) | 53 (47/7) | 1 | 0.52 | 1 | 0.74 |
|  | Second | 56 (41/2) | 51 (45/9) | 1.49<br>(0.44-5.01) |  | 1.20<br>(0.71-2.02) |  |
|  | Third or more | 10 (7/3) | 7 (6/4) | 1.79<br>(0.63-2.16) |  | 0.92<br>(0.33-2.58) |  |
| **Pregnancy trimesters** | First | 39 (28/7) | 30 (27) | 1 | 0.37 | 1 | 0.56 |
|  | Second | 50 (36/7) | 48 (43/2) | 0/78<br>(0/36-1/69) |  | 1.24<br>(0.67-2.31) |  |
|  | Third | 47 (34/6) | 33 (29/8) | 1/27<br>(0/68-2/36) |  | 0.91<br>(0.47-1.75) |  |
| **Age** | ≤ 30 | 88 (64/7) | 65 (58/5) | 1 | 0.37 | 1 | 0.32 |
|  | > 30 | 48 (35/3) | 46 (41/5) | 1.38<br>0.67-2.82 |  | 1.29<br>(0.77-2.17) |  |

## Discussion

HAV and HEV are the leading causes of acute self-limiting viral hepatitis worldwide. The severe form of infection can result in a significant mortality rate in pregnant women. Our study showed that the overall seroprevalence of HEV infection in pregnant women’s study population was 0.8%. Our finding, similar to the rate of HEV in the developed countries, was lower than the seroprevalence of HEV infection among pregnant women in other developing countries, such as India (33.6%), Sudan (41%), and Egypt (84.3%) [11, 12]. Moreover, some studies in Spain reported that the rate of HEV-positive IgG was 3.6% [13]. These differences could be explained by the difference in socioeconomic and educational status, hygiene level, and the endemic status of the virus.

Our results also showed that the overall seroprevalence of HAV infection in the study population for pregnant women was 44.9%, almost similar to a previous study by Farajzadegan et al. in which HAV seroprevalence in Iran was reported to be 51% [14]. The seroprevalence of HAV varies worldwide. In developed countries, health and vaccination have reduced the incidence of HAV [15]. In Asia, the seroprevalence of HAV varies between high, moderate, and low levels in different regions. It is noteworthy that Iran has been considered an HAV endemic region in recent world reports [16].

We found a significant inverse association between educational status and HAV because HAV-positive women’s rate increased with lower education levels (p = 0.003), which was consistent with a previous study by Ramezani et al. [17]. No significant relationship between age and frequency of HAV was observed due to the proximity of age groups in the present study, although our results showed that the anti-HAV antibodies were more detected in people aged ≤ 30 years than that of other age groups. These results were consistent with the results of a study by Mohebbi et al. [18].

While the rate of infection among rural residents was higher than that of urban residents, living in a city or village does not significantly affect the HAV infection rate. The reason for this can be attributed to access to safe drinking water, use of a hygienic toilet system, increasing awareness of the general population, and more sanitation in these areas. It is of interest to mention a strong confounding bias in comparing this stratification factor. Many people now residing in cities lived in the village for many years, especially in their childhood. Therefore, they might have been infected with the HAV in the past. Thus, these patients’ past medical history should also be considered, which was possible in this study.

Our results also showed that HAV incidence is higher among women in their second trimester, followed by third and first trimesters, respectively. Nonetheless, no significant relationship was observed in this regard that might be attributable to the small sample size.

## Conclusion

Due to improved health conditions, the risk of developing hepatitis is reduced in childhood but increased in adulthood. Also, it is crucial to consider the many complications of these diseases during pregnancy. Therefore, the detection of hepatitis A and E viruses in pregnant women is important. Thus, ways to promote health problems, such as increasing the awareness of these infections and vaccination in the general population, can prevent adverse complications.

## Data Availability

The data that support the findings of this study are available from the corresponding author upon reasonable request.

## Funding statement

This study was fully supported by the vice-chancellor for research and technology of Babol University of Medical Sciences.

### Acknowledgments

The authors thank the Department of Infectious diseases of Babol University of Medical Sciences.

## Ethics approval statement

This study protocol was approved by the ethics committee of Babol University of Medical Sciences (MUBABOL.HRI.REC.1396.141).

